# A Co-Creation Process Toward Sustainable Adoption of Integrated Care for Prevention of Unplanned Hospitalizations

**DOI:** 10.1101/2023.08.03.23293537

**Authors:** Carmen Herranz, Alba Gómez, Carme Hernández, Rubèn González-Colom, Joan Carles Contel, Isaac Cano, Jordi Piera-Jiménez, Josep Roca

**Affiliations:** Consorci Sanitari de Salut de Barcelona Esquerra (CAPSBE). Barcelona, ES; Research and Innovation Management, Hospital Clínic de Barcelona. Barcelona ES.; Fundació de Recerca Clínic Barcelona - Institut d’Investigació Biomèdica August Pi I Sunyer (FRCB-IDIBAPS), Barcelona, ES; Centro de Investigación Biomédica en Red de Enfermedades Respiratorias (CIBERES). Madrid, ES; Catalan Health Department. Barcelona, ES; Digitalization for the Sustainability of the Healthcare System DS3-IDIBELL. L’Hospitalet de Llobregat, ES; Faculty of Informatics, Multimedia and Telecommunications, Universitat Oberta de Catalunya. Barcelona, ES; Catalan Health Service. Barcelona (CatSalut), ES; Hospital Clínic de Barcelona (HCB). Barcelona, ES; Universitat de Barcelona (UB). Barcelona, ES

**Keywords:** Adaptive Case Management, Co-creation, Design Thinking, Digitalization, Prevention of Unplanned Hospital Admissions

## Abstract

**Introduction:** Complex chronic patients (CCP) are prone to unplanned hospitalizations leading to a high burden on healthcare systems. To date, interventions to prevent unplanned admissions show inconclusive results. We report a co-creation process performed into the EU initiative JADECARE (2020-2023) to elaborate an integrated care program aiming at preventing unplanned hospitalizations.

**Methods:** A two-phase process of structured interviews and design thinking (DT) sessions was conducted. Firstly, we assessed the management of CCP in Catalonia (ES) through twenty interviews (five patients and fifteen professionals), including the results of a cluster analysis of 761 hospitalizations, followed by two DT sessions (Oct 2021 to Feb 2022). Then, we examined the 30- and 90-day post-discharge periods of 49,604 hospitalizations as input for two DT sessions with seven professionals.

**Discussion:** The co-creation process identified poor personalization of the interventions, the need for organizational changes, immature digitalization, and suboptimal services evaluation as main explanatory factors of the observed efficacy-effectiveness gap. Additionally, a program for prevention of unplanned hospitalizations, to be evaluated during 2023-2025, was generated.

**Conclusions:** A digitally enabled adaptive case management approach to foster collaborative work, as well as organizational re-engineering, are endorsed for value-based prevention of unplanned hospitalizations.

## INTRODUCTION

Complex chronic patients (CCP) (1) frequently face unplanned hospitalizations, either due to episodes of exacerbation, or during periods associated with increased vulnerability, like the transition from hospital to the community after hospital discharge (2). It is widely accepted that unplanned hospital admissions generate significant dysfunctions, and a high financial burden, on healthcare systems worldwide (3).

The reports from reputed international organizations (4,5) indicate that a relevant percentage of unplanned hospitalizations are avoidable. However, the existing literature examining interventions to reduce admissions in selected CCP with episodes of severe exacerbations (6) and in transitional care programs (7) aiming at decreasing early readmissions after hospital discharge present inconclusive findings This can be attributed to the heterogeneity of the interventions and/or suboptimal description of the protocols leading to poor comparability, among other factors. Overall, there is a clear consensus on the need to explore the potential of clinical processes to efficiently prevent unplanned hospitalizations within a care continuum scenario (8).

To this end, a co-creation process was undertaken within the Catalan original Good Practice (oGP) of the European Union (EU) Joint Action JADECARE (9), running in the period 2020-2023. Two factors triggered the initiative of the case practice. Firstly, during the pre-implementation phase of JADECARE, internal discussions within the Catalan oGP highlighted the necessity of reevaluating the preventive management of CCP. The second triggering factor was the unsolved efficacy-effectiveness gap (EEG) observed between the promising results of a two-centre randomized controlled trial (RCT), carried out in 2006 (10) and the lack of effectiveness observed when the same intervention was replicated in a pragmatic RCT conducted during 2015 (11).

The case practice focused on designing and adopting person-centred integrated care interventions for selected CCP to prevent unplanned hospitalizations. This approach covered two situations: firstly, selected patients showing a high risk of severe exacerbations due to clinical severity, poor self-management and/or environmental factors; and secondly, transitional care strategies after hospital discharge.

Ultimately, the targeted objectives in this case practice are: i) To identify key factors explaining the observed EEG; ii) To produce well-defined interventions, including identification of target candidates, as well as the steps needed to achieve mainstream adoption, and iii) To evaluate the potential for generalization/transferability of such interventions at the EU level.

The report sequentially describes the regional context of the analysis, the main achievements of the two-phase co-creation process based on structured interviews and DT sessions, and the process for the elaboration of the program for the prevention of unplanned hospitalizations to be tested during 2023-2025.

## DEVELOPMENT AND IMPLEMENTATION OF THE CASE PRACTICE

### Setting the scene

Since 1981, the Catalan Health System (12,13) has been fully responsible for policy formulation, organization management, service delivery, and financial aspects of the health services in Catalonia (ES), with 7.7 million citizens. While key areas such as basic legislation, interregional coordination, pharmaceutical policy, international health policy, and educational requirements remain under the Spanish central government’s control, the Catalan Health System holds complete jurisdiction over subsidiary legislation, public health initiatives, the organizational structure of the healthcare system, accreditation, and planning processes, as well as purchasing and service provision activities.

The regional system ensures universal coverage and free access at the point of use. Healthcare expenditure represents approximately 8% of the gross domestic product paid by taxes (14). The single public payer, CatSalut (15), has a well-defined separation from a network of multiple providers fully accounted for in terms of health objectives, activity, invoicing system, financial aspects, and evaluation system. Basic principles and traits of the regional health system are: i) civil society participation, ii) access equity, and iii) robust tradition of regional health planning. The latter has an essential role in triggering key sustainable changes at the regional level, as recently assessed by the 2020 WHO report on the “*Thirty-year Retrospective of Catalan Health Planning: Driver of Health System Transformation”* (16). It is of note that the level of citizen’s satisfaction regarding health services, measured yearly through the Health Survey of Catalonia (ESCA) - “Enquesta de Salut de Catalunya” (17), is on average 8/10.

### Integrated Care in Catalonia (ES)

– The regional health planning (18,19) has played a backbone role in triggering the health system transformations toward digitally supported integrated health and social care services during 2011-2020.

Since the early 1980s, the primary healthcare centres (PHCs) in Catalonia have been the first point of contact for individuals seeking healthcare services. PHCs are staffed by general practitioners, community nurses, paediatricians, and other healthcare professionals. Citizens are assigned to a specific PHC based on their residence, such that each PHC is responsible for serving a defined population within its catchment area. This structure, designed to be patient-centred, plays a major role in facilitating the care continuum. The second major component in the integrated care scenario is the different modalities of services, grouped into the concept of intermediate care and end-of-life services. Such area was conceived in 1986 under the seminal program “Vida als Anys” (Life to Years) (20) and further evolved toward a myriad of interconnected service modalities, displayed in **Table 1**. During 2011-2020, coordination of social support services, dependent on city councils and healthcare, has also been a core objective.

**Table 1.**
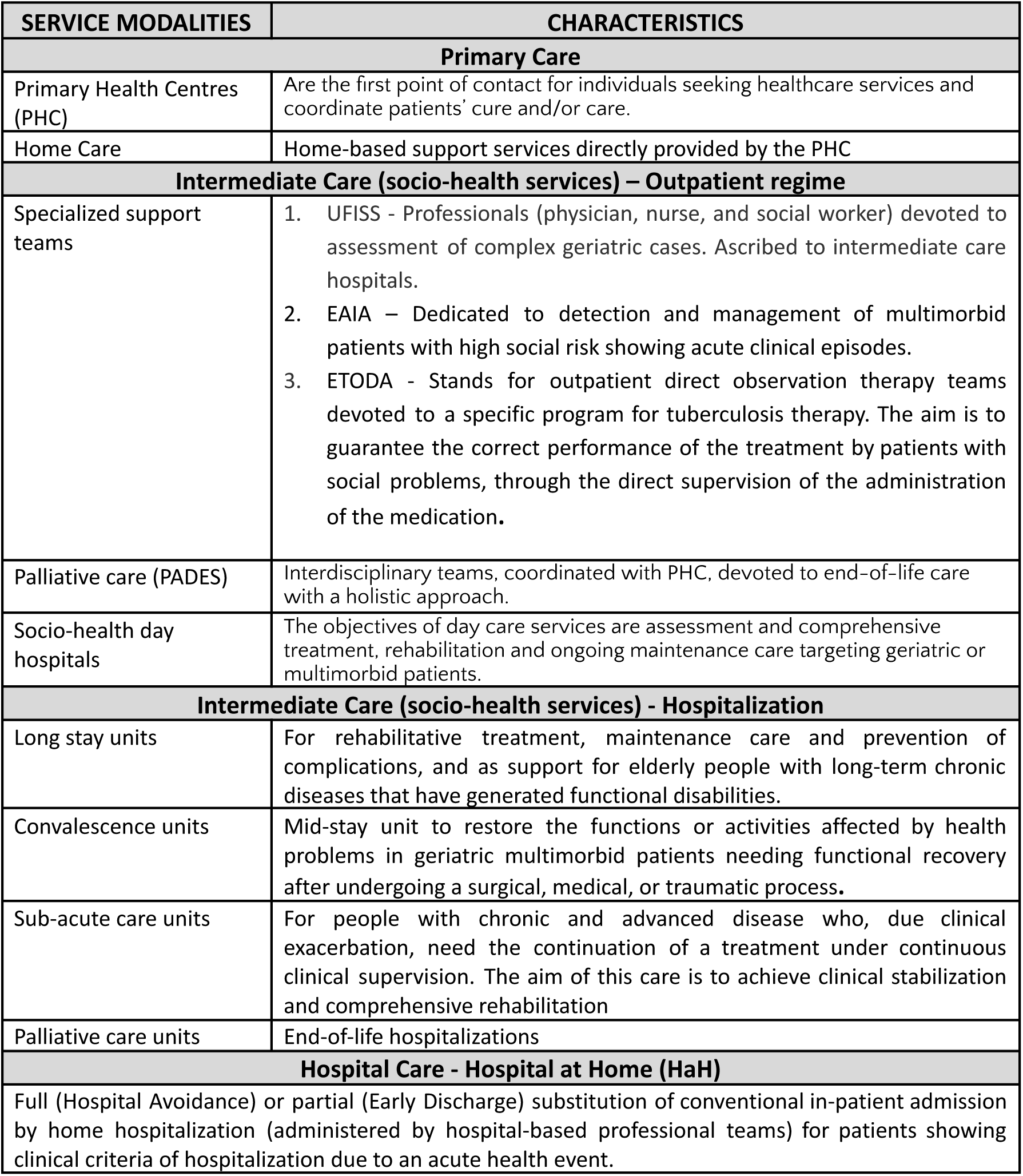
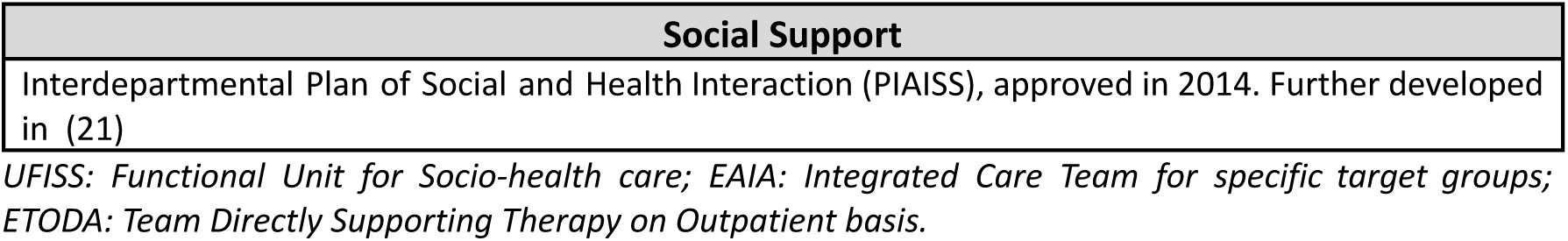
Service modalities supporting care continuum in Catalonia.

Two distinct documents influenced the development of the current integrated care case, each contributing to its conceptual framework. One document encompasses the fundamental characteristics of the regional care model for chronic patients (1), providing operational definitions of frailty, CCP, and advanced chronic patients (ACPs) with a life expectancy of less than 18 months. The second report outlines the ongoing digital strategies for 2021-2025 (22), specifically focusing on implementing a knowledge-based digital platform to support cloud-based health services.

### Co-creation process within JADECARE

The co-creation process was conducted following a grounded theory approach, combining inputs from recent quantitative analyses (23,24) and DT methodologies (25), as indicated in **Figure 1**, wherein the timeline and the focus of each of the two phases are depicted. The first co-creation phase, named “Discovery”, had a twofold purpose: i) to identify actionable factors to enhance regional CCP management; and ii) to generate specific recommendations to be tested during 2023-2025. Whereas the second phase, named “Confirmation”, took advantage of a comprehensive analysis of Hospital a Home (HaH) in Catalonia (ES), executed during the last semester of 2022. Such analysis provided the opportunity to assess the post-discharge period in a large dataset of hospital admissions. The aim was to confirm some assumptions and proposals generated in the Discovery phase.

**Figure 1.**
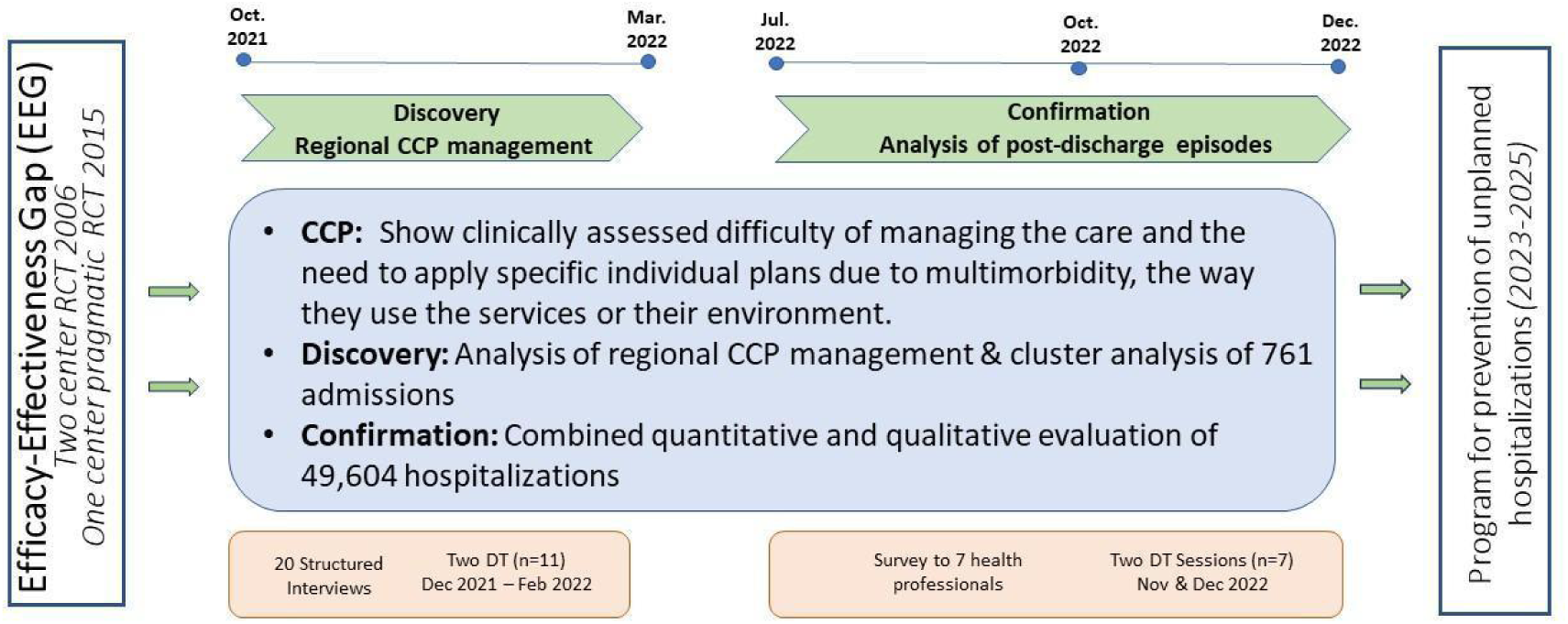
Two-phase co-creation process timeline. - **A trigger:** The efficacy-effectiveness gap (EEG) seen between two studies carried out in 2006 (10) and 2015 (11). **The Discovery phase**: devoted to the analysis of regional Complex Chronic Patients (CCP) management and identifying the main explicable factors for the EEG. CCP represent 4% of the population, allocated above P95 of the population-based risk stratification pyramid. **The Confirmation phase**, assessing value generation of Hospital at Home, was used to analyse the interactions between the hospital teams and community-based services, reflecting the status of vertical integration (24). **The final outcome** is the elaboration of a program for preventing unplanned hospitalizations to be tested during 2023-2024. DT: Design Thinking. RCT: Randomized controlled trial

Overall, the recommendations produced during the two co-creation phases were the basis for elaborating the roadmap for deploying and adopting a program for the prevention of unplanned hospitalizations in selected CCP.

For the entire co-creation process, the research team was assisted by a qualitative research and service design specialist recruited as a facilitator for planning and leading the expert panel discussions. A description of the methodological aspects, including the selection criteria of the participants in the structured interviews and the DT sessions and the results of the process, is provided in the online **Supplementary material**.

Discovery Phase: The regional complex chronic patients’ program. The process was initiated with twenty structured interviews, mean duration 45 min each, to selected representative individuals, followed by two DT sessions of 120 min duration each. The interviews were administered to five representative patients and fifteen leading health professionals (**Table 1S**).

For the patients, the interviews were structured to capture the reported experience on the following aspects: i) initiation of the acute episode, ii) emergency room admission, iii) hospitalization period; iv) follow-up after hospital discharge, v) intermediate care, if needed, vi) home-based support and services, if needed; vii) primary care services; and viii) care continuum.

For the fifteen leading professionals, the interview addressed the following items relative to CCP: i) problems associated to the identification of CCP, ii) roles and interactions among healthcare professionals in the care of CCP, iii) satisfactoriness of the current CCP management, iv) challenges, unmet actionable needs and proposals for CCP management, v) proposals to reduce unplanned hospitalizations in CCP, and vi) challenges, unmet actionable needs and proposals to enhanced transitional care after hospital discharge.

Eleven leading health professionals with different profiles (nine of them also participating in the interviews) attended the two DT sessions (**Table 1S**).

The results of the interviews were processed and analysed to nourish the first DT session. The information gathered was used to generate a Context Analysis (**Figure 1S**), whereas the patients’ answers were the basis for elaborating an Empathy Map (**Figure 2S**).

The second item used as input data for the first DT session was the asymmetrical results of the studies conducted in 2006 and 2015 (10,11) showing the EEG alluded to above.

Finally, the last input was a recent study conducted at Hospital Clinic Barcelona (HCB) (23), combining predictive modelling for mortality and early re-admissions after hospital discharge and a cluster analysis in 761 patients. The study outcomes triggered a lively debate on tailored interventions to improve the personalization of transitional care after hospital discharge. **Figure 2**, reproduced from (23), displays the main traits of the four clusters of patients identified in the study, as well as their mortality rate and use of healthcare resources after discharge. Clusters #1 and #2 include patients with a mean age in the early seventies. It is of note that patients in cluster #2, with potentially reversible risk factors related to unhealthy lifestyles, clearly showed worse health status and poorer prognosis than those in cluster #1. Whereas individuals in clusters #3 and #4 showed a similar mean age in the mid-eighties, with high medical or social complexity, respectively. The study’s results fostered the debate on service selection depending on the patient profile, for example, home care or intermediate care programs (Clusters #3 and #4) versus community-based prevention of unplanned hospitalizations (Clusters #1 and #2).

**Figure 2 legend.**
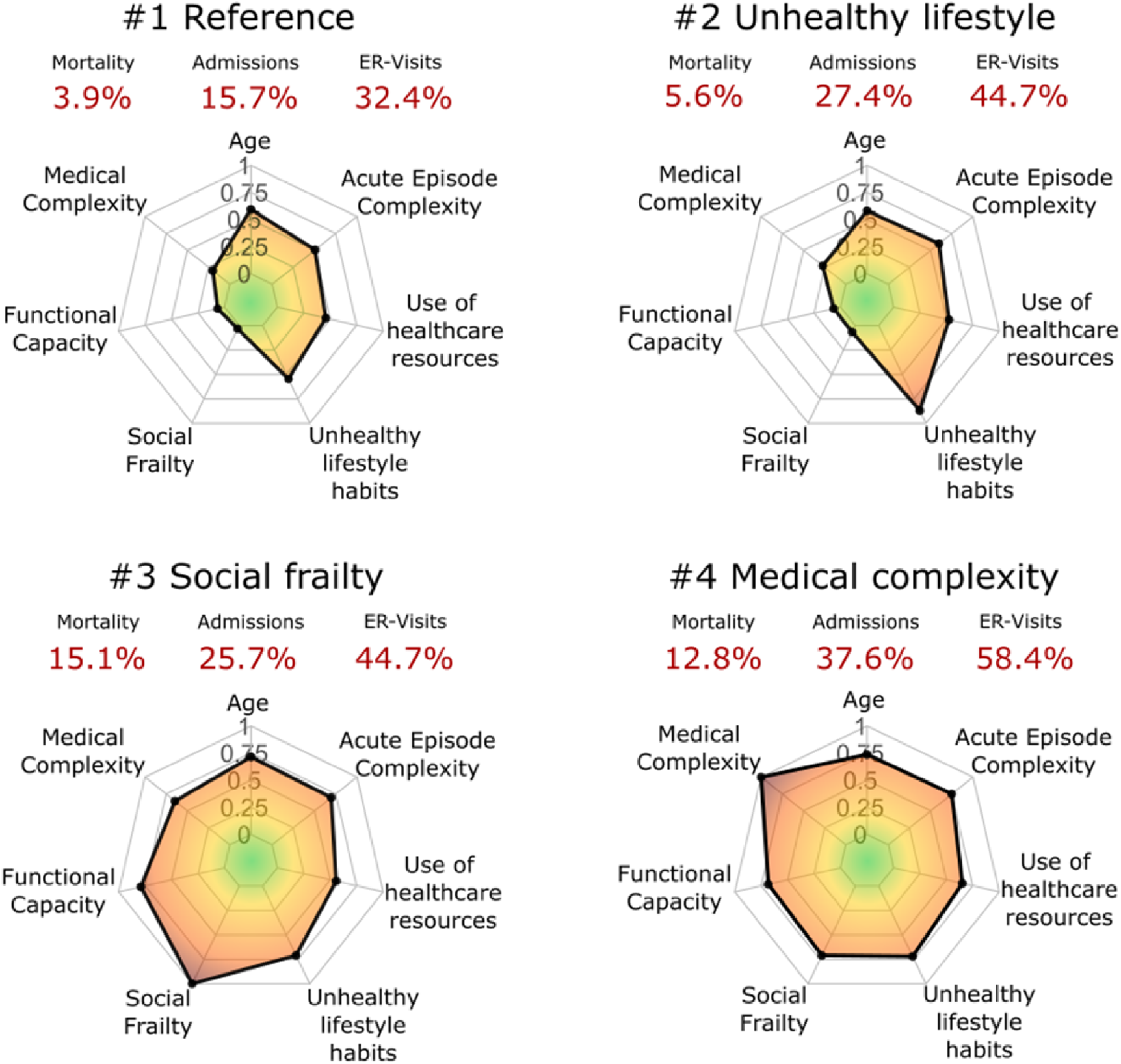
– Radar plots of the main characteristics of the four clusters of patients identified in (23). All the features are normalised and grouped into seven categories: i) age; ii) medical complexity; iii) functional capacity; iv) social frailty; v) unhealthy lifestyle habits; vi) use of healthcare resources; and vii) acute episode complexity. The mortality rates, hospital admissions and emergency room visits are displayed in red.

The first DT session was devoted to analysing the characteristics of the current regional CCP management. At the end of the first session, we performed a SWOT analysis on CCP management, as well as a summary of the results of the first DT session (**Figure 3S**), that were used to feed the second session focused on exploring actions to improve CCP outcomes within a care continuum scenario. An Impact-Feasibility Matrix (**Figure 4S)** was generated from the debate.

The Discovery phase’s main results identified four actionable EEG determinants to be analysed further in the Confirmation Phase. The results of the Discovery phase support the hypothesis that an efficient program to prevent unplanned hospitalizations in selected CCP could be elaborated by properly articulating the following areas:

*1. Change management* was identified as the most relevant determinant of the EEG requiring attention for action. However, several different factors/needs were considered under the epigraph, namely: i) a better definition of the boundaries and interplay among different modalities of services depicted in **Table 1**, with emphasis on integration between health and social care services, ii) a definition of professionals’ profiles and roles required for enhanced management of selected CCP to be included in a program for prevention of unplanned hospitalizations, iii) unmet specific needs in terms of education and training of those professionals, iv) request for a deeper debate defining patients’ profiles, including their specifics needs for cure and/or care, and v) shared care agreements across healthcare tiers and collaborative work are not adequately supported.
*2. Personalization of the interventions –* The cluster analysis results (**Figure 2**) (23) show four different patients’ profiles with specific needs for personalization of integrated care strategies. These results reinforced the need for exploring the boundaries and interplay among different modalities of clinical and social care services indicated in **Table 1**, as well as the debate on cure and/or care needs in specific groups of CCP. Moreover, the personalization in terms of the type of concurrent diseases, and disease severity, focuses on the complementarities between disease-focused *vs* patient-oriented management. Also, case management strategies to monitor the progress of the patient’s status should be considered. These aspects highlight the importance of carefully selecting services and dynamically personalizing interventions over time. Moreover, such requirements have also impact on the roles of the professionals involved in the preventive program.
*3. Mature Digitalization –* Current digital support was considered not to meet the requirements for the management of CCP. The group identified the following requirements to provide scalable digitalization: i) support collaborative work with an adaptive case management (ACM) approach. By implementing ACM, integrated care processes can be designed, developed, and implemented in a way that empowers patients, clinicians, and other stakeholders, leading to improved health outcomes and better patient experiences., ii) supporting cloud-based digital health tools with an ACM approach, interoperable (ad-hoc) with existing electronic health records from different suppliers across healthcare tiers.
*4. Adoption and assessment –* The transition from evidence-based efficacy of any intervention to the demonstration of effectiveness and healthcare value generation assessed using a Triple or Quadruple Aim approach (26) was considered a must. Nevertheless, the applicability of the current assessment tools in real-world scenarios shows major limitations. As reported in (27) there is an urgent need to develop and validate applicable tools to evaluate PROMs/PREMs, or their surrogates, for mainstream interventions in real-world scenarios. Also, identifying relevant key performance indicators (KPI) and elaborating appropriate dashboards for quality assurance after service adoption were proposed as core requirements. To this end, refinement of the comprehensive evaluation approach described in (28), aiming at enhancing its applicability, was strongly recommended.

### Confirmation Phase: Analysis of Post-Discharge Episodes

The assessment of value generation, and transferability, of HaH in Catalonia (ES) during the period 2015-2019 (24), carried out in the context of JADECARE, offered the opportunity to analyse a large dataset of post-discharge episodes. Also, to explore the interactions between the hospital teams and community-based services, reflecting the status of vertical integration.

Comprehensive quantitative and qualitative analyses were conducted on all episodes of HaH, and their corresponding control group of patients under conventional hospital admissions, generated through propensity score matching techniques. A total of 49,604 episodes from 27 different hospitals were examined. The analysis included patients’ characteristics, multimorbidity-complexity, use of healthcare resources and expenditure during four well-defined periods: i) one year before the acute episode, ii) during the acute episode requiring hospital admission, iii) 30-days, and iv) 90-days after hospital discharge. A large portion of the patients fell into the spectrum of CCP, as reported in (24), with no differences in health outcomes between modalities of hospitalization, either HaH or in-hospital admissions.

The outcomes of the quantitative analysis of the hospital admission episodes, and the results of a survey previously administered to a panel of seven experts in HaH, fed the debates of the two subsequent DT sessions participated by the same group of 7 experts (**Figure 1**, **Table 2S**). While both quantitative and qualitative analyses of value generation of HaH were reported in (24), the assessment of the post-discharge period, including the interactions between hospital and community-based professionals during the transitional care period after discharge, was essentially confirmatory of the considerations raised in the Discovery phase.

Interestingly, the study showed huge heterogeneities among healthcare suppliers within the same healthcare system. The analysis of the factors behind such heterogeneities fully supported the need to consider the four actionable identified during the Discovery phase. Moreover, the two DT sessions indicated a high potential of the HaH teams to foster productive interactions between specialized professionals and community-based teams leading, to mature vertical integration.

### Program for prevention of unplanned hospitalizations

The research team at HCB-IDIBAPS assumed the recommendations generated in the two-phase co-creation process (**Figure 1**) to elaborate the basis for a program aiming at preventing unplanned hospitalizations in selected CCP, described as follows.

The program’s main aims of are: i) early detection and management of exacerbations, as well as early identification of undesirable events after hospital discharge, ii) patients’ empowerment for self-management, and iii) shared care agreements and collaborative work across levels of care.

The profile of the target candidates for the program should fulfil the following traits: i) multimorbid outpatients with a high risk for hospital admissions because of a previous history of repeated severe exacerbations requiring admissions, ii) patients with a high morbidity burden, measured with the Adjusted Morbidity Groups (AMG) (29,30) scoring, that is, all those cases allocated in the at-risk stratum of the population risk pyramid of Catalonia (i.e. above percentile 80 of the regional AMG scoring distribution) and iii) patients not being allocated to home care or intermediate care programs depicted in **Table 1**.

The AMG score is an aggregative index based on diagnostic codes, which indicates the burden of an individual’s morbid conditions through a disease-specific weighting deduced from statistical analysis based on mortality and the utilization of health services. Moreover, all hospital post-discharges with an AMG score above percentile 80 should be evaluated as potential candidates for personalized 30-days transitional care interventions.

The research team proposes three sequential phases to finetune and evaluate the program to prevent unplanned hospitalizations, facilitating its adoption as a mainstream service at the end of a two-year (2023–2025) process, including: i) Preliminary Studies, ii) Assessment of preventive interventions, and iii) Adoption as a mainstream service.

During the two years, the testing will be done in a cohort of 200 multimorbid outpatients showing two main traits: i) multimorbid individual allocated close to the peak of the Catalan risk stratification pyramid (> P_80_) based on the AMG scoring (29,30), and ii) having chronic obstructive pulmonary disorders of moderate to severe intensity as one of the main diagnoses. Such a cohort will be followed up for a longer period, as part of the EU project, “Knowledge for improving indoor air quality and health” (K-Health in Air) (31), aiming to explore the relationships between indoor air quality and health status in chronic patients.

**Table 2** indicates the main actions, directly derived from the co-creation process, to be evaluated and subsequently incorporated into the mainstream program to prevent unplanned admissions in selected CCP.

**Table 2.** Actions to be included in the program for prevention of unplanned hospitalizations.

| ACTIONS | COMMENTS |
| --- | --- |
| **1. CHANGE MANAGEMENT** |  |
| 1.1. Define flexible clinical processes with a holistic approach. | Elaborate the framework of the clinical process considering: i) clinical endpoints, actions associated to main diagnosis and co-morbidities, as well as environmental and social factors. |
| 1.2. Define roles and profile of the advanced care nurse | Key roles are: i) patients' empowerment for self-management, ii) care and cure actions following plans defined in 1.1, iii) early detection/management of exacerbations. Double ascription to primary care & Hospital at Home teams. Coordination with intermediate care service modalities |
| 1.3. Integrate primary to quaternary preventions | Evolution from current focus on primary prevention to integration of all prevention levels in the management of the program candidates |
| 1.4. Redefine interactions between nurse and patient | Initial motivational intervention followed by patient's agreements on a personalized care plan (non-pharmacological interventions). Patient's activation and empowerment following the procedures reported in (32,33) |
| 1.5. Training programs for professionals and patients | Education/training of professionals and patients before the program initiation following the innovative approaches reported. |
| **2. PERSONALIZATION OF THE INTERVENTIONS** |  |
| 2.1 Service selection | Selection of the patient as candidate to the program or allocation into other service modalities indicated in **Table 1** |
| 2.2. Harmonize disease- vs patient-oriented approaches. | The intervention (personalization of the clinical process) must consider the individual disease (type, severity, and progress), as well as their potential interactions (also regarding pharmacological aspects). |
| 2.3. Consider social and environmental factors | The holistic approach of the intervention requires consideration of the non-clinical aspects (social status, education level, environment, etc...) that may have influence on health status |
| 2.4. Consider evolution of health status overtime | The characteristics of the intervention decided in the initial evaluation requires adaptation to the progress of the patient's condition. The balance between cure and care should be considered. |
| **3. MATURE DIGITAL SUPPORT** |  |
| 3.1. Adaptive Case Management (ACM) | Combinations of multiple factors influencing the patient's health status, and unexpected events (exacerbations), requires flexible management of the clinical process which can be achieved with an ACM approach. |
| 3.2. Communication Channel. | A communication channel based on chat (including intelligent chatbots) with multimedia support is essential to provide proactive interactions among stakeholders. |
| 3.3. Collaborative work | Digital support to collaborative work among professionals and with patients across healthcare tiers is a must for executing shared care agreements. |
| 3.4. Capture of patient's self-tracking data | Efficient patients' input data from: i) non-disruptive sensors, ii) chat with the advanced care nurse, and/or iii) short questionnaires (with Likert scales) may play a relevant role in decision making and knowledge generation. |
| 3.5. Ad-hoc integration | Cloud-based technologies with ad-hoc integration with providers' health information systems is the proposed approach for provision of digital support to the service. |
| **4. APPLICABLE ASSESSMENT IN REAL-WORLD SETTINGS** |  |
| 4.1. Evaluation of the process of implementation | Evolution of classical tools for assessment of the service deployment (i.e. Consolidated Framework for Implementation Research, CFIR (34)) must be adopted to enhance applicability in real-world settings. |
| 4.2. (PROMS)/ (PREMS) | Role of sensor measurements (i.e., Heart Rate Variability), info from the chat and short questions (Likert scale) are candidates to substitute classical questionnaires (33,35–37). |
| 4.3. User-profiled dashboards | Identification of Key Performance Indicators (KPI) and build-up management dashboards contribute to service quality assurance over time. |

The initial six months, from October 2023 to March 2024, will be devoted to the evaluation of innovative strategies for patient assessment consisting of three parallel independent study protocols addressing early detection and enhanced management of acute episodes of exacerbations. The three items to be explored are, respectively: i) feasibility and impact of continuously monitoring the heart rate variability, as compared with the periodic administration of standard health questionnaires (diaries) (38), ii) the feasibility of pragmatically capturing Patient Reported Outcomes/Patient Reported Experiences (PROMS/PREMS) using mobile technology (33,35,37,39); and iii) the role of forced oscillation technique (FOT), as compared to forced spirometry (FS) (40).

The subsequent phase will consist of personalized preventive interventions, following the recommendations indicated in **Table 2**. The interventions will be run by a nurse case manager over twelve months, from April 2024 to March 2025. The nurse will have a double assignment to the primary care teams and the HaH team at HCB. The digital support to the intervention will be based on a customized version of the cloud-based ACM platform named Health Circuit™ (33). Evaluation of the effectiveness and value generation of the intervention using a Quadruple Aim approach is planned, as well as the assessment of the deployment process with a pragmatic modification of the Consolidated Framework for Implementation Research (CFIR) approach (34). The identification of Key Performance Indicators (KPI), and elaboration of a dashboard, for subsequent continuous quality assurance and management of the intervention, will be made.

The final six months, April to September 2025, will be used to adapt of the profiles of the target candidates and the program’s characteristics to the requirements of mainstream service. This period will also be employed to consolidate the final steps of certification of the digital tools supporting the service.

## DISCUSSION

The current integrated care case took advantage of the lessons learnt throughout the regional adoption of digitally supported integrated care guided by the two Catalan Health Plans 2011-2015 and 2016-2020 (18,19).

JADECARE (2020–2023) (9) has provided an adequate frame for critically evaluating the achievements and failures over more than one decade of adoption. Also, has prompted several quantitative assessments (23,41) that have contributed to the co-creation process.

The Discovery phase allowed a structured analysis of the unmet needs of the regional management of CCP and facilitated the identification of four actionable areas for improvement, tackling: i) organizational aspects and novel professional roles, ii) personalization of the services, iii) innovations in digital support fostering collaborative work and use of ACM principle, and iv) structured support to the assessment of the transition from a study protocol to a sustainable adoption of the intervention. It is of note that the outcomes of the Confirmation phase were fully aligned with those previously identified in the Discovery phase, confirming the role of HaH as a relevant facilitator of vertical integration.

Beyond the recommendations alluded to above, the case practice has activated the application of specific technological and organizational solutions, generating synergies with early phases of the EU project K-Health in Air (31) as a testing field of the program for the prevention of unplanned hospitalizations during 2023-2025. It is reasonably expected that, at the end of the two-year period, the finetuned interventions will be ready for adoption as a mainstream service in a real-world scenario.

The authors are cognizant of the fact that certain intricacies of the organizational solutions presented in the document may need site-specific adaptations when contemplating their applicability to other European scenarios that exhibit varying health system structures.

To our knowledge, the current study illustrates all the steps of the long journey from the necessary demonstration of evidence-based efficacy with RCT, to a sustainable adoption of the intervention, that is: i) need for assessment of effectiveness, ii) evaluation of the deployment process using implementation science methodologies, ii) assessment of healthcare value generation, and iv) identification of KPI (and elaboration of user-profiled dashboards) for continuous quality assurance of the service after adoption. Since those steps are expected for most integrated care interventions, we believe that the recommendations raised by the current study are generalizable to other use cases beyond the program for preventing unplanned hospitalizations.

## LESSONS LEARNED

The two-phase co-creation process has generated the following key learnings:

1. **A nurse case manager in the community**, can play a central role for enhanced management of CCP, also bridging with hospital-based specialists, as well as with other modalities of clinical and social care services for chronic patients.
2. **Personalization of the interventions** aiming at preventing unplanned hospitalizations according to: i) the evolving patient’s health status, ii) the relative weight of the different patient’s morbidities, as well as iii) to other determinants of frailty and complexity, constitutes a key requirement.
3. **Innovative cloud-based digital support**, following ACM principles, and interoperable with different health and social care suppliers, fosters more informed decisions through collaborative work and facilitates end-used engagement.
4. **The underwent co-creation process has demonstrated its effectiveness** as a systematic and robust pathway for identifying the key elements necessary for the intervention’s transition from demonstrating evidence-based efficacy to being adopted as a value-based mainstream service. Those steps, reported in the case practice, are generalizable to other integrated care services.
5. **Transferability of recommendations, for the four actionable areas, at European level is possible.** But the need for site specific organizational adjustments shall be considered according to the characteristics of each health system.

## CONCLUSIONS

The present study outlines the co-creation process employed to identify four crucial factors (change management, intervention personalization, advanced digitalization, and improved real-world service evaluation) influencing the observed EEG in previous studies that aimed to test an intervention for preventing unplanned hospitalizations in CCP.

Based on those main determinants, the study provides recommendations for a scalable program for preventing unplanned hospitalizations targeting selected CCP and transitional care strategies after hospital discharge. Moreover, the research proposes the sequential steps to pave the way towards the adoption of the intervention as a mainstream value-based service.

## ETHICAL APPROVAL

The quantitative studies displayed in the discovery phase received approval from the Ethical Committee for Human Research at the Hospital Clínic of Barcelona in 2017 (Approval numbers: 26/04/2017, 2017-0451, and 2017-0452). Whereas the studies presented during the confirmation phase obtained approval from the Ethical Committee for Human Research at Hospital Clinic de Barcelona in 2021 (Approval number: HCB/2021/0768). In both instances, the Ethical Committee for Human Research at Hospital Clinic de Barcelona waived the requirement for informed consent to collect, analyze, and publish retrospectively acquired and fully anonymized data for these non-interventional studies.

All data handling adhered to the General Data Protection Regulation 2016/679 guidelines, which govern data protection and privacy for all individuals within the European Union. The study was conducted in strict accordance with the relevant legal requirements of biomedical research (Biomedical Research Act 14/2007 of July 3).

The co-creation process involved the participation from five patients who voluntarily granted informed consent to participate in the study and disseminate its results.

## Data Availability

All data produced in the present study are available upon reasonable request to the authors

## ACKNOWLEDGMENTS

This research was funded by JADECARE project-HP-JA-2019 - Grant Agreement n° 951442 a European Union’s Health Program 2014-2020.

The authors thank David Font, Joan Escarrabill and Gemma Yago from Hospital Clínic de Barcelona; Luís González-de Paz and Antoni Sisó from the Consorci d’Atenció Primaria de Salud de Barcelona Esquerra; Esther Limón and Juan Jose Zamora from Institut Català de la Salut; Jordi Piera from Catalan Health Service; and Marco Inzitari from Parc Sanitari Pere Virgili for their valuable participation in interviews and Discovery phase; Carme Hernández and Jose Antonio Rodríguez from Hospital Clínic de Barcelona; Susanna Torres and Zoe Herreras from the Consorci d’Atenció Primaria de Salud de Barcelona Esquerra; Joan Cales Contel from Ministry of Catalan Health; and Oscar Solans from Catalan Health Service for their participation in interviews; and Jordi Amblas and Belen Enfedaque from Ministry of Catalan Health for their important participation in Discovery phase.

The authors also thank David Nicolás from Hospital Clínic de Barcelona; Francesc Xavier Jiménez from Hospital Vall d’Hebron; Montserrat Suárez and Elvira Torné from Catalan Health Service; Eulalia Villegas-Bruguera from Hospital Dos de Maig; Gerard Carot from Digitalization for the Sustainability of the Healthcare (DS3)-IDIBELL; and Mireia Espallargues from Agència de Qualitat i Avaluació Sanitàries de Catalunya for their valuable collaboration in the Confirmation phase.

We appreciate the time and efforts of all patients who provided information. We are indebted to Fernando Ozores (BuenaIdea) for his excellent assistance in the interviews and leading the DT sessions.

## ACRONYMS

ACM: Adaptive Case Management
ACPs: Advanced Chronic Patients
AMG: Adjusted Morbidity Groups
ATDOM: Home Care
CCP: Complex chronic patients
CFIR: Consolidated Framework for Implementation Research
DT: Design-thinking
EAIA: Integrated Care Team for specific target groups
EEG: Efficacy-effectiveness gap
ESCA: Enquesta de Salut de Catalunya
ETODA: Team Directly Supporting Therapy on Outpatient basis.
EU: European Union
DT: Design Thinking
FOT: Forced Oscillation Technique
FS: Forced Spirometry
HaH: Hospital Care - Hospital at Home
HCB: Hospital Clinic Barcelona
JADECARE: The Joint Action on implementation of digitally enabled integrated person-centred care.
KPI: Key performance indicators
PADES: Palliative care
PHCs: Primary healthcare centres
PIAISS: Plan of Social and Health Interaction
RCT: Randomized controlled trial
UFISS: Functional Unit for Socio-health care

