## Supplementary material for "A Co-Creation Process Toward Sustainable Adoption of Integrated Care for Prevention of Unplanned Hospitalizations": Online supplement

*Herranz C. et al. 2023*

*Online supplementary material*

Discovery phase

### Table 1S – Participants in the Discovery Phase: interviews and/or design thinking sessions.

| **Name** | **Expertise & Position** | **Filiation** | **Participation** |
| --- | --- | --- | --- |
| Zoe Herreras | General Practitioner (GP) & Head of Care Processes | CAPSBE | I only |
| Antoni Sisó | GP & CAMFIC president | CAPSBE | I + DT |
| Susanna Torres | Social worker | CAPSBE | I only |
| Luis González | Nurse & Technician | CAPSBE | I + DT |
| Juan José Zamora | Nurse & Head of nursing processes | ICS | I + DT |
| Esther Limón | GP & Education coordinator & CAMFIC treasurer | ICS | I+DT |
| Gemma Yago | Advanced practice nurse at the Diabetes Unit | HCB | I+DT |
| David Font | MD & Strategy and Planning Director | HCB | I+ DT |
| Jose Antonio Rodríguez | Nurse & Coordinator Geriatric Unit | HCB | I only |
| Carmen Hernandez | Nurse & Innovation Unit | HCB | I only |
| Joan Escarrabill | MD & Director of Chronic Care | HCB | I+DT |
| Marco Inzitari | MD & Director of Intermediate Care | Parc Sanitari Pere Virgili | I + DT |
| Jordi Piera | Computer Engineer & Director of the Digital Health Strategy Office. CatSalut | Ministry of Health | I+ DT |
| Joan Carles Contel | Nurse & Staff member of the Chronic Care Program | Ministry of Health | I only |
| Oscar Solans | MD & Functional manager of eHealth | CatSalut | I only |
| Jordi Ambàs | Director of the Integrated Care Agency | Ministry of Health | DT only |
| Belen Enfedaque | Deputy director of primary and community care | Ministry of Health | DT only |

*GP: General Practitioner; CAPSBE: Primary Care Consortium at Barcelona-Esquerra; I: Interview; CAMFIC: Catalan Society of Primary Care; DT: Design Thinking sessions; ICS: Institut Catalan of Health; MD: Medical Doctor; CatSalut: Catalan Health Services is the single-public payer.*

### Interviews and Design thinking (DT) sessions

During the months of October to December 2021, in-depth interviews were conducted with 20 key informants, including 5 patients and 15 clinical professionals and healthcare service managers, chosen for their leadership in the Catalan Health System.

The interviews with the patients focused on their experiences in the care process. For the other key informants, the interviews addressed topics such as the identification of patients with complex needs (CCP), the role of healthcare professionals, current management of CCP, and proposals to improve care and reduce readmissions. The interviews were carried out by the first author (CH) and a qualitative research and service design specialist (FO, Buenaidea). On average, the interviews lasted for 45 minutes each.

*Patients’ recruitment* - A total of 25 patients were selected from the community, CAPSBE (Primary Care Consortium of Barcelona-Esquerra) as potential candidates for the interviews. The inclusion criteria were: i) At least one hospital admission in the last year, ii) Two or more chronic pathologies and iii) Adjusted Morbidity Group (AMG) Score 3-4, which corresponds to > P80 in the Catalan risk stratification pyramid. We excluded those patients presenting: i) Difficulties in conducting an interview (sensory problems, cognitive impairments, or severe mental illness), and ii) Refusing to participate. The final five patients, in whom the interviews were conducted, were selected balancing the following criteria to achieve the highest representativeness: i) Male/female, ii) With/without caregiver and iii) With transition from hospital to home/hospital-intermediate-care-home.

Professionals’ recruitment (**Table 1S**)– Fifteen experts were selected based on their leadership in integrated care, as well as the coverage of three areas: i) Clinical activities (physicians, nurses, and social workers), ii) Managers and policy makers (micro-, meso-, and macro levels), and iii) Technological expertise. Representativeness of different healthcare tiers (Primary care, Hospital care, Intermediate care, and Macro-management) was also considered.

Two design thinking (DT) sessions were conducted with fifteen key professionals (**Table 1S**) between January and February 2022. All fifteen professionals participating in the interviews were invited to participate in the DT sessions, but only nine of them accepted. The absences of the remaining six were due to scheduling conflict. Moreover, two additional leaders could attend only the DT sessions. Such that the final number of attendees to the two DT session was 11.

For the first DT session, a Context analysis (**Figure 1S**) and an Empathy map (**Figure 2S**) were prepared, based on the information gathered from the interviews. Moreover, input from quantitative studies (1-3), as reported in the main document, were included in the debate. Discussion and consensus-building took place regarding the ideas presented, and proposals were collected. The results of the first DT session were used to generate a SWOT analysis and to prepare a summary of the results of the session (**Figure 3S**). Both exercises were input material for the second DT session of the Discovery phase.

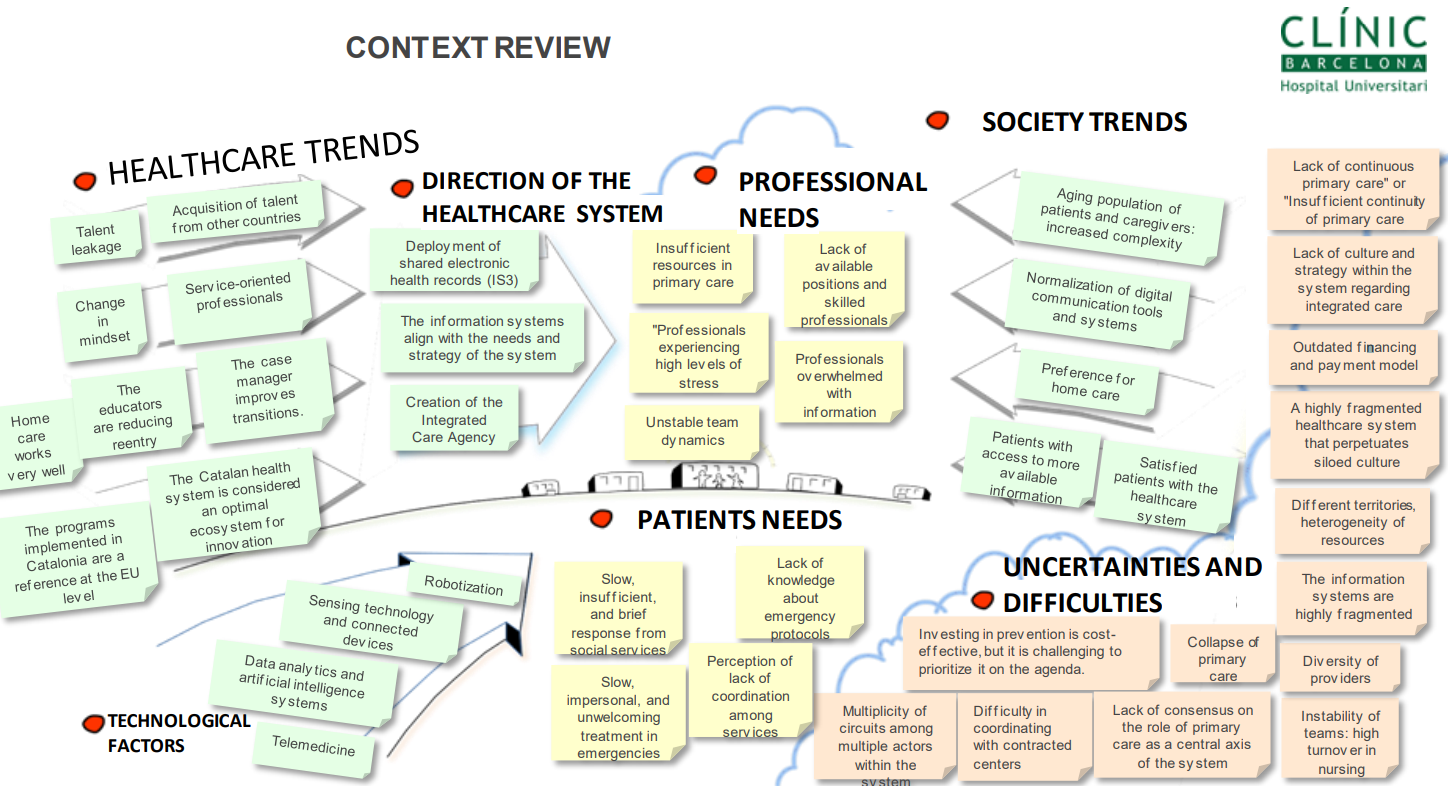

***Figure 1S. Context Analysis****. General understanding of the patient’s and experts’ ideas collected during the interviews, aligned with seven different topics related to healthcare implementation.*

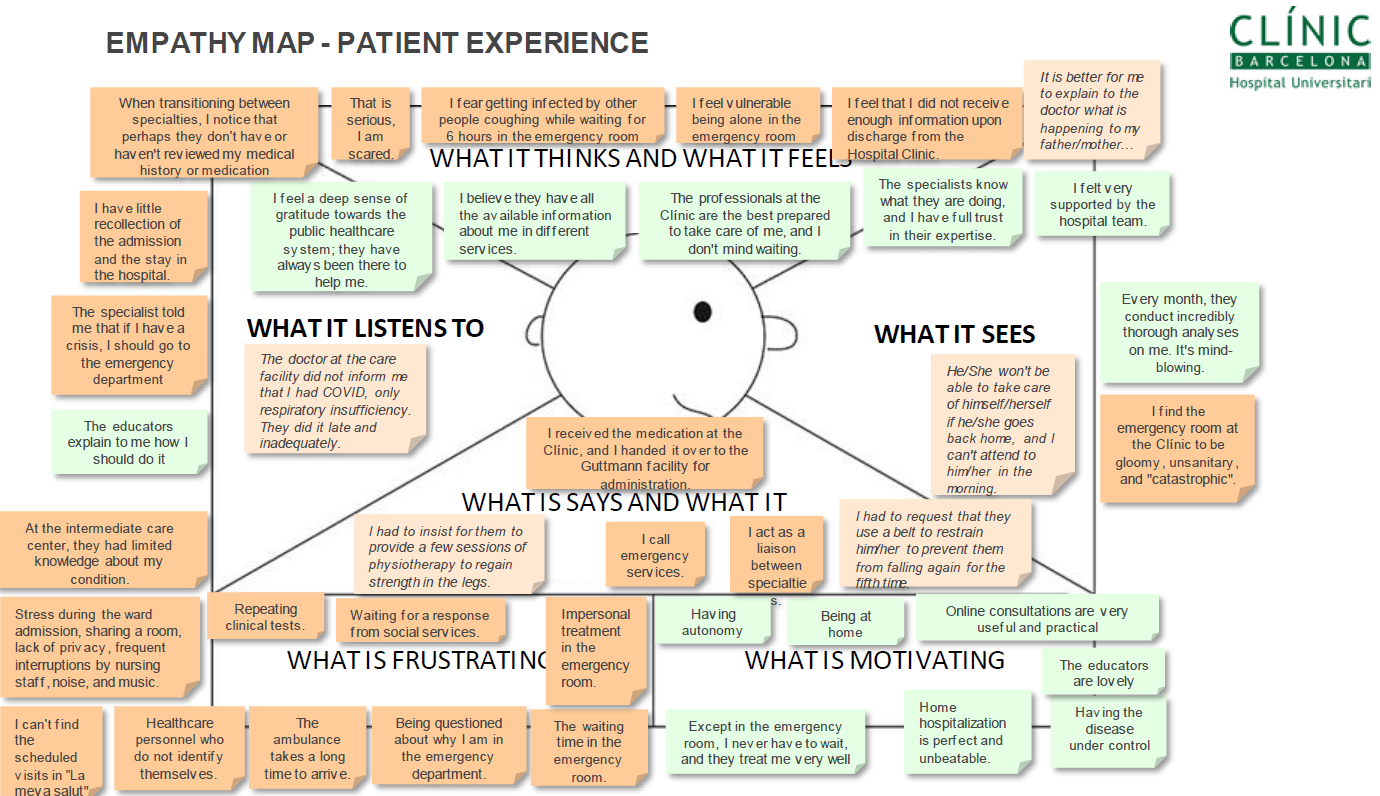

***Figure 2S****.* ***Empathy Map****. Summary of the patients' interviews. Focused on their experience in the care process, ranging from the onset of the episode to the stay and discharge in intermediate care. Four areas that are confined to patient perception, segregated by whether they are* *discerned as a frustrating or motivating situation.*

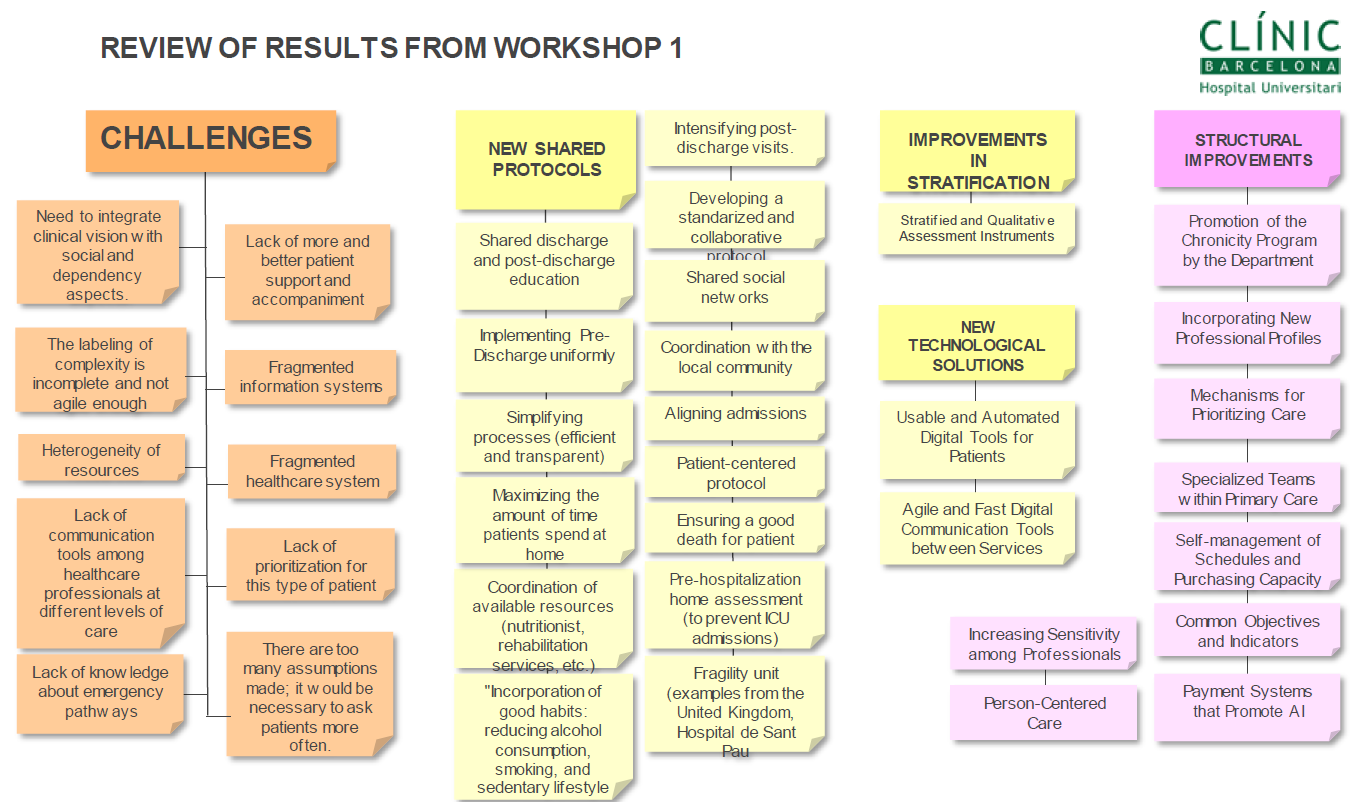

***Figure 3S. Results from Workshop 1****. Key challenges and proposals gathered during the first DT design session were most relevant to the participants.*

During the second DT session, all the participants from the previous session remained engaged. Building upon the information gathered and the variables explored previously, they delved deeper into the challenges and proposals at hand. During the second DT session, a context analysis and an Impact feasibility matrix was constructed (**Figure 4S**).

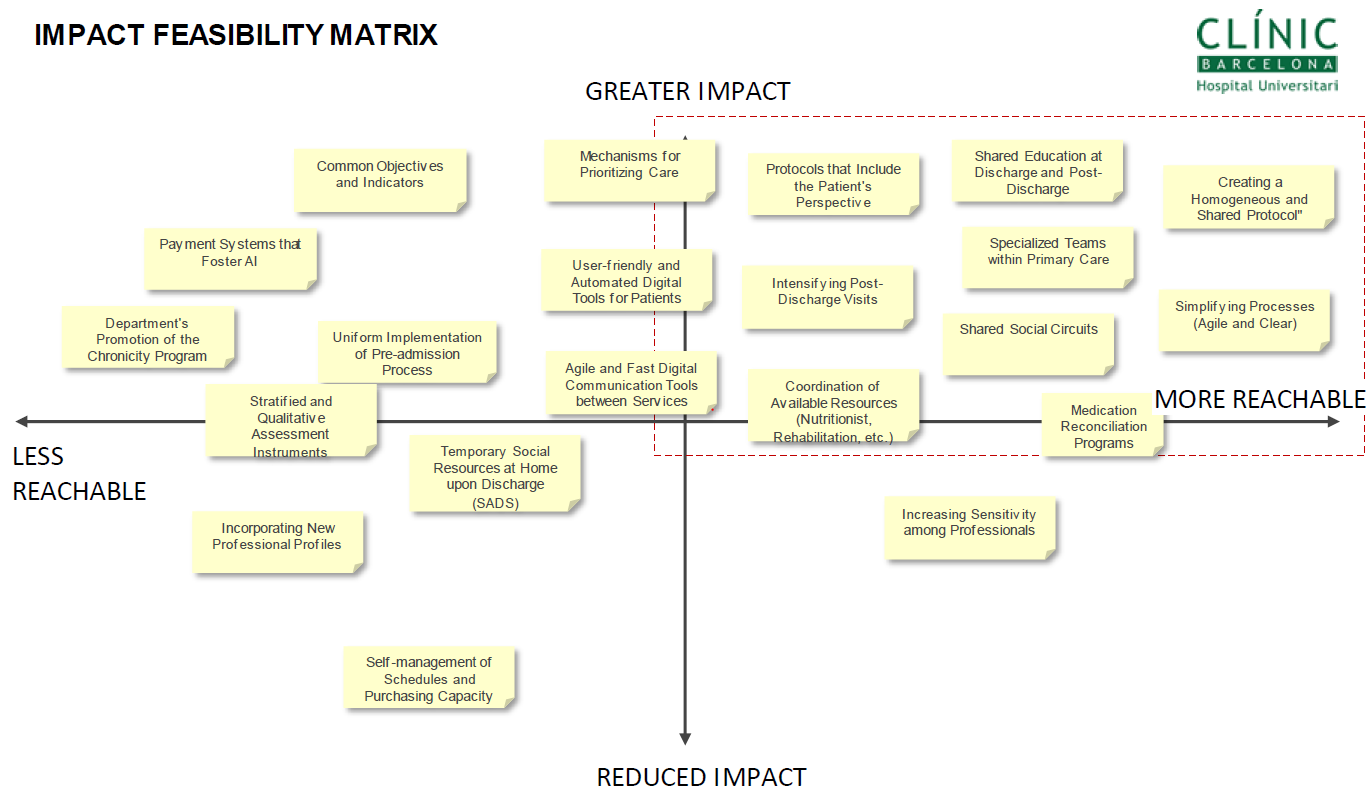

***Figure 4S. Impact feasibility matrix.*** *This illustration was generated during the second DT session with the most important variables detected. A discussion was held regarding the operational aspects of the proposed solutions. The Impact feasibility matrix* *was enhanced for better understanding and making informed decisions, allowing us to identify areas of advantage, areas that need improvement, potential opportunities to leverage, and potential outcomes to mitigate.*

Codes, subthemes, and themes were generated using Atlas.ti 9 software.

Confirmation phase

### Table 2S. Participants in the Confirmation phase: Survey (S) and Design Thinking (DT) sessions

| **Name** | **Expertise & Position** | **Filiation** | **Participation** |
| --- | --- | --- | --- |
| David Nicolas | MD & Coordinator of the HaH | HCB | S + DT |
| Francesc Xavier Jiménez | MD & Coordinator of the HaH | Hosp. Vall d’Hebron | S + DT |
| Eulalia Villegas-Bruguera | MD & Coordinator of the HaH | Hosp. Dos de Maig | S + DT |
| Carme Hernandez | Nurse. PhD & Innovation Unit & Former HaH coordinator | HCB | S + DT |
| Mireia Espallargues | Staff member (HaH specialist) | AQuAS | S + DT |
| Montserrat Suárez | Staff member (HaH specialist) | CatSalut | S+ DT |
| Elvira Torné | Staff member (HaH specialist) | CatSalut | S + DT |

*MD: Medical Doctor; HaH: Hospital at Home, HCB: Hospital Clinic de Barcelona; S: Survey; DT: Two Design Thinking sessions; AQuAS: Catalan Health Quality Agency; CatSalut: Catalan Health Services is the single-public payer.*

The panel of 7 experts included 1-to-2 representatives of the most relevant organizations in implementing or assessing HaH services in Catalonia: two members from the Catalan-Balearic Society of Hospital at Home (FXJ and EV-B), two staff members from the Catalan Health Service (MS and ET), one staff member from the Health Quality and Assessment Agency of Catalonia (AQuAS) (ME), and two HaH experts from the local JADECARE team (DN and CH). Four out of the seven experts were clinical leaders of different HaH programs. A qualitative research and service design specialist (F.O., BuenaIdea) was recruited as a facilitator for planning and leading the expert panel discussions.

### Survey and Design thinking sessions

Overall, the experts agreed that HaH is safe and provides value to the healthcare system, with similar health outcomes than conventional hospitalization, and positive impacts on patients’ and professionals’ experience. HaH may also result in savings associated with fewer personnel and structure requirements. The experts agreed that heterogeneity in patient profile and outcomes was expected and identified three important sources of this heterogeneity: i) maturity of HaH teams (i.e., mature teams tend to admit older and more complex patients), ii) hospital strategies to use HaH in a sub-set of patients with specific diagnoses, and iii) local ecosystem (e.g., lack or availability of certain integrated care services in the area). The high potential shown by HaH to foster vertical integration was unanimously endorsed by the expert group. The full set of results of the quantitative and qualitative analyses have been reported in (4).
